# Respiratory virus infection dynamics and genomic surveillance to detect seasonal influenza subtypes in wastewater: a longitudinal study in Bengaluru, India

**DOI:** 10.1101/2025.01.13.25320458

**Authors:** Namrta Daroch, KK Subash, Vishwanath Srikantaiah, Rakesh Mishra, Farah Ishtiaq

**Author notes:** Corresponding author: Farah Ishtiaq.

## Abstract

Recent global pandemics have been caused by respiratory viruses found in both humans and animals with zoonotic spillover potential. Many respiratory viruses, such as severe acute respiratory syndrome coronavirus 2 (SARS-CoV-2), respiratory syncytial virus (RSV), influenza A virus (IAV), and influenza B virus (IBV), have overlapping ecology and share similar symptoms. However, respiratory disease surveillance is often passive and relies on testing clinical specimens. Wastewater surveillance has been widely used as an epidemiological tool for early detection of SARS-CoV-2 variants and can differentiate between respiratory virus infections and SARS-CoV-2 peaks at a community level. In this retrospective longitudinal study covering three SARS-CoV-2 Omicron waves, we conducted a monthly sampling for 28 months (812 samples) between August 2021 and December 2023 at 28 sewershed sites in Bengaluru city (∼11 million inhabitants). Using RT-qPCR kits, we quantified SARS-CoV-2 RNA concentrations, IAV, IBV and RSV to understand community viral infection occurrences. We provide evidence of changes in the relative abundance of influenza subtypes and SARS-CoV-2 variants driving emerging peaks in wastewater data. We found 86% of samples positive for SARS-CoV-2, while the positivity rates for influenza virus and RSV were lower (37% for IAV, 16 % for IBV, and 15% for RSV). We observed an increase in influenza viruses in the post monsoon season (August to December) and remained present in January and February, while being practically absent the rest of the year. RSV showed a similar trend to IAV, with comparable quantities. Furthermore, we identified all eight segments of influenza virus genomes and emerging SARS-CoV-2 variants in wastewater samples. Wastewater surveillance provides data on the abundance of respiratory viruses in urban Bengaluru that would not be reported otherwise. Under the One Health framework, wastewater surveillance can offer early warning signs and enhance the traceability of infectious diseases in both wildlife and humans.

## 1. Introduction

Acute respiratory illnesses (ARI) pose a major healthcare challenge in developing countries. In India, ARI is a leading cause of mortality in children under five with substantial geographical disparities [1,2]. Enhanced policies and programmes are needed to reduce vaccine-preventable deaths and achieve the Sustainable Development Goals child survival targets by 2030 [2]. Additionally, many respiratory viruses, such as SARS-CoV-2, respiratory syncytial virus (RSV), and influenza, share similar symptoms, hindering clinical diagnosis [3,4]. Given the overlapping host-virus ecology, co-infection of SARS-CoV-2 and influenza viruses in humans could alter disease transmission patterns and aggravate disease burden [5].

Influenza viruses belong to the Orthomyxoviridae family and are divided into types A, B, and C. Influenza is a highly contagious viral disease caused by multiple strains influenza virus. Influenza A virus strains are classified into subtypes by the combination of the two proteins; Haemagglutinin (HA) and Neuraminidase (NA), found on the outside of the virus. Influenza viruses undergo antigenic evolution through antigenic drift and shift in their surface glycoproteins. This necessitates frequent updates of influenza vaccine components and timely surveillance [6].

Of the 16 HA subtypes that have been identified in birds, 5 HA subtypes of IAVs (H5N1, H5N6, H5N8, H6N1, H7N9, H7N2, H7N3, H7N7, H9N2, H10N7, and H10N8) are known to cause human infections. The H1N1 and H3N2 IAVs are continuing to circulate in humans as seasonal influenza. In addition to humans, IAVs have a wide range of host animal species in nature, especially wild aquatic birds (e.g., Anseriformes and Charadriiformes), the reservoir hosts of IAVs and mild disease in poultry such as breathing problems and reduced egg production. Birds can be infected with low pathogenic avian influenza virus (LPAI: H1-H16) and the IAV isolated from or adapted to avian hosts are avian influenza viruses (AIV). Among these, the H5 subtype has a nearly global distribution in birds. High pathogenic avian influenza virus (HPAI: only H5 and H7 subtypes) causes outbreaks and associated mortality in poultry and wild birds [7]. HPAI, subtype H5N1, constitutes one of the world’s most important health and economic concerns given the catastrophic impact of epizootics on the poultry industry, the high mortality attending spill over in humans, and its potential as a source subtype for a future pandemic.

In India, respiratory disease surveillance is negligible or biased due to underreporting. The Integrated Disease Surveillance Programme (IDSP) under the National Centre for Disease Control is responsible for monitoring the situation weekly to detect early warning signals of impending outbreaks or changes in the epidemiology of diseases of public health importance and respond rapidly. As per IDSP data, India experiences bimodal influenza seasons every year: one from January to March and other in post monsoon season [https://pib.gov.in/PressReleasePage.aspx?PRID=1905602]. RSV infections are observed during monsoons, which peaks during September and October months [8–9]. There is a need for a complementary surveillance approach independent of individual testing to identify spatial and temporal patterns in co-circulating respiratory viruses. Wastewater surveillance from selected locations to detect, quantify, and characterize infectious disease pathogens is cost-effective and particularly useful in low-resource settings where obtaining representative data on the infectious disease burden is challenging [10].

Densely populated cities play a huge role in the spread of emerging diseases, particularly those transmitted via respiratory and faecal-oral routes. Since August 2021, our team represents a collaboration between researchers from Bangalore Life Science Cluster (BLiSc) and the city utility responsible for sewerage have been running a citywide wastewater surveillance of SARS-CoV-2 in Bengaluru city. Through this surveillance program, wastewater samples from the influent of 28 sewage treatment plants (STP), covering ∼11 million inhabitants, have reported strong relationships between RNA from SARS-CoV-2 found in wastewater and rates of COVID-19 cases in the community [11] and characterized the spatiotemporal variation in antibiotic resistome and corresponding bacterial community [12].

In response to growing need to differentiate between SARS-CoV-2 outbreaks from seasonal respiratory viruses to zoonotic viruses, we aimed to test the use of wastewater surveillance to evaluate and compare RNA levels of IAV/IBV, RSV-A/RSV-B, and SARS-CoV-2 in wastewater in Bengaluru. We expect these results would complement influenza surveillance system (IDSP) and help in defining testing priorities and vaccination drives. This proof-of-concept study can be used in devising guidelines for monitoring human and animal respiratory viruses. Overall, this study lays the groundwork for systematic characterization of viral diversity in wastewater that can potentially influence public health decision-making.

## 2. Materials and Methods

### 2.1 Wastewater sample collection

We sampled influent wastewater from 28 centralized sewage treatment plants (STPs) in Bengaluru, India. Grab samples were collected once a month from each STP from August 2021-December 2023 between 0800-1400 h. These STPs covered different catchment area sizes: eight small (serving 10,000 − 60,000 population), nine medium (serving 100,000 – 350,000 population), and nine large (serving 400,000 − 2,480,000 population) STPs (Fig. 1 A). The details of inflow rate, STP capacity (volume of water), population size is provided in Supplementary Table 1.

**Fig. 1:**
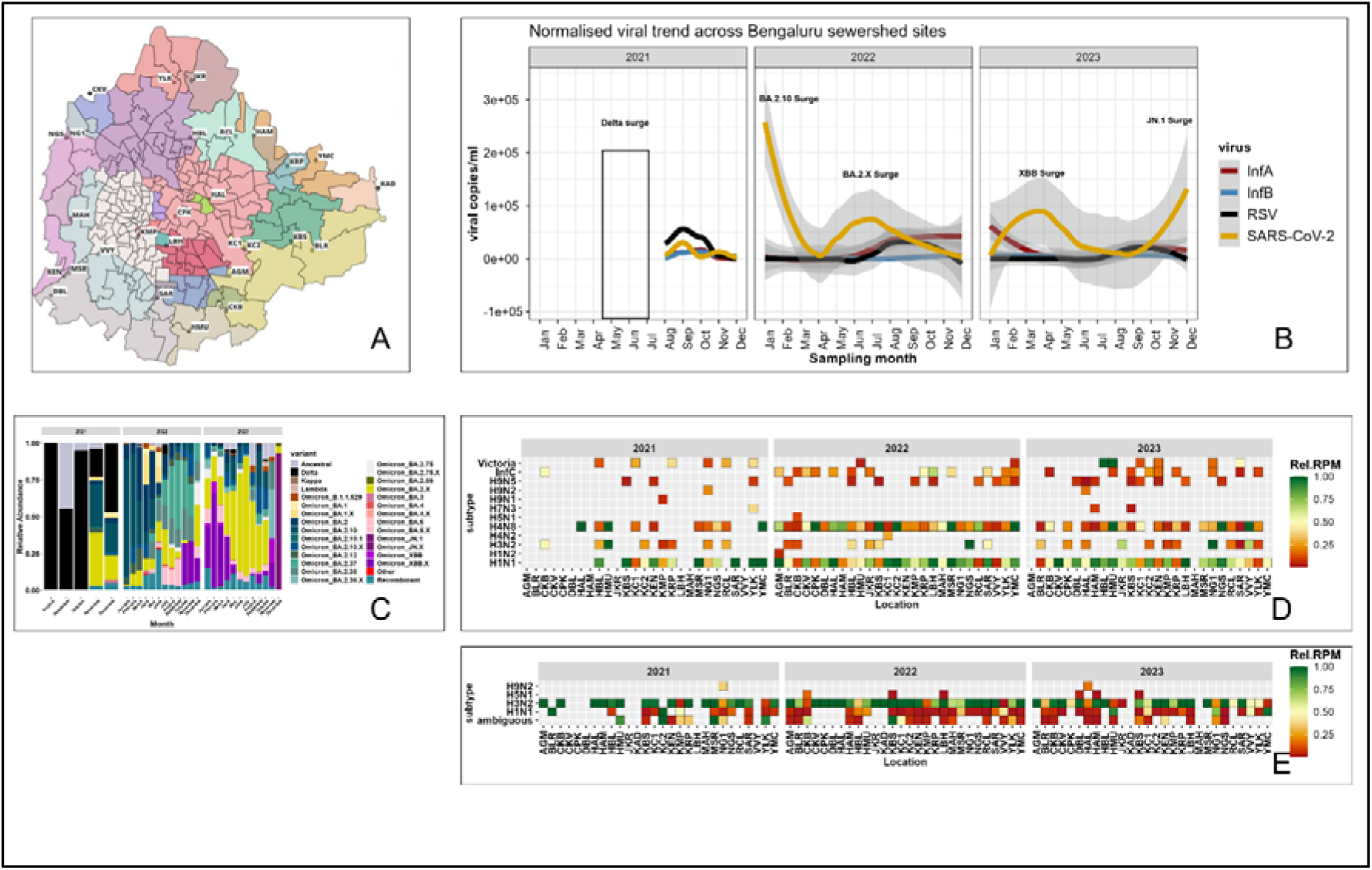
Map of Bengaluru showing geographical location of 28 sewershed sites screened for respiratory viruses from August 2021-December 2023. Fig. 1B: Temporal trends of respiratory viruses in non-treated wastewater sampled. Fig. 1C: SARS-CoV-2 variant dynamics in wastewater. Fig. 1D: Influenza detection and subtype assignment using the RefSeq genomes of influenza viruses (A/B/C) revealed diversity of influenza A (H1N1, H1N2, H3N2, H4N2, H5N1, H7N3, H9N1, H9N2, H9N5), influenza B (Victoria) and influenza C (InfC) subtypes across sewershed sites. Fig. 1E: Influenza detection and subtype assignment using iav_serotype pipeline revealed H1N1, H3N2, H5N1 and H9N2 subtypes.

Samples were collected in 200 mL plastic bottles, tightly sealed upon collection, and stored at 4°C in the field. All samples were processed within 24 h at a biosafety level 2 facility as described previously [11]. Briefly, samples were subjected to heat inactivation and incubated in a 200 mL bottle at 60°C for 90 min and divided into three replicates. Subsequently, 40 mL of master sample (total 120 mL from 200 mL bottle) was transferred to three 50 mL centrifuge tubes containing 0.9 g NaCl and 4 g polyethylene glycol (PEG; 8000 MW). The PEG and NaCl mixture in the samples was vortexed until it dissolved, and the solution was centrifuged at 11000 rpm for 30 min at 4°C. After discarding the supernatant, 600 µL of lysis buffer was added to resuspend the pellets. Finally, the solution was transferred to a centrifuge tube and briefly vortexed to dissolve the pellet. Viral RNA was extracted using Qiagen Viral RNA mini kit protocol following the manufacturer’s instructions and 50 µL of elution was stored at -80°C until subsequent analysis.

### 2.2. Viral Reverse Transcriptase-quantitative PCR (RT-qPCR)

We used two RT-qPCR kits designed to screen SARS-CoV-2, RSV and Influenza viruses in wastewater. For SARS-COV-2, *Gene*Path Dx CoViDx One v2.1.1TK-Quantitative multiplex RT-qPCR kit was used. The kit targets three viral genes: N-gene, RdRp-gene, and E-gene along with a human control gene (RNAase P gene) [11]. The kit is designed for a quantitative analysis (positive or negative), COVID-19 Viral Load Calculation Tool (RUO; https://coviquant.genepathdx.com/) was used for the quantifying viral load in the samples.

For viral load calculation, three standards provided within the kit were used: high standard = 5000 copies/μL, medium standard = 500 copies/μL and low standard = 50 copies/μL. The online tool uses the Ct values (cut-off 35) of high, medium and low and calculate the unknown in copies/μl of each sample.

For generic detection of RSV-A/B (M gene), Influenza-A (M gene), and Influenza-B (non-structural gene), the *GenePath* Dx Multiplex with a human control gene (RNase P gene), was used. The test can detect H1N1 and H3N2 of Influenza A/Victoria and Influenza B/Yamagata strains. However, the kit does not distinguish between IAV and IBV or their subtypes. Therefore, to quantify the IAV and IBV in wastewater, RT-qPCR was carried out using CDC FluSC2 assay (https://archive.cdc.gov/www_cdc_gov/coronavirus/2019-ncov/lab/multiplex.html) with a total 15 μl volume containing 5 μl of eluted RNA and 3.75 μl of Luna Probe One-Step RT-qPCR 4X Mix with UDG (NEB, Catalog no: M3019L) under the following cycling conditions: 25 °C for 30 s, 55°C for 10min, 95 °C for 1min, and 45 cycles of 95 °C for 10 s, then 56 °C for 1min on a QuantStudio™ 5 Real-Time PCR System. The fluorescent signal was measured during the annealing step. Primer sequences and concentration of primers and probes used are available in Supplemental material 1. Samples were run in duplicate wells and included negative extraction control and no template control. Samples were considered positive if Ct values were ≤35.

To determine copy numbers, of IAV (M-gene) and IBV (NS-gene) a standard curve of the linearized pUC57 plasmid was to determine the genomic copy numbers (See Supplementary material 1 for details). RNA extracts of negative and invalid samples were tested using Pepper mottle virus (PMMoV) as a measure of indicators of human fecal contamination.

### 2.3 Viral load modelling

We used consolidated data for each month to estimate change in viral concentrations. We calculated the daily viral load, *VL* (copies/d) for SARS-CoV-2, influenza A and influenza B, and RSV by normalizing raw viral load, *C* (copies/mL) to the average daily STP flow, *Q(L/d)* using equation 1 [13] and *P* is the population of inhabitants the catchment of the respective sewershed site.

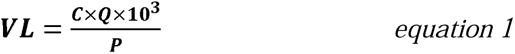

We used Kendall’s tau (*τ*) statistic [14] to estimate the association between viral concentration. Values close to 1 indicate strong agreement, and values close to -1 indicate strong disagreement. This test was selected as the data were not normally distributed. We performed six pairwise tests, and tau values were considered significantly different from zero when *p*<0.00009.

### 2.4 Genomic surveillance

#### (i) Metagenomic shotgun sequencing for Influenza

An unbiased metagenomic approach was used to genetically characterise pan-viral diversity [15]. Here we analysed data for influenza viruses and results of other viruses will be reported elsewhere. A subset of 242 grab samples, with 10 samples for each month positive and negative for influenza, were selected for sequencing at the Next Generation Genomics Facility in BLiSc. RNA extraction was quantified using a Qubit™ Fluorometer (Invitrogen) with a Qubit HS RNA assay kit. RNA integrity was analyzed using a 4200 Tape Station (Agilent) and an RNA Screen Tape assay kit. To enrich the viral RNA content before library preparation, ribosomal RNA (rRNA) was depleted using a combination of two kits: NEBNext® rRNA Depletion Kit v2 (Human/Mouse/Rat) (Cat No. NEB E7405) and NEBNext® rRNA Depletion Kit (Bacteria) (Cat No. NEB E7860), along with RNA Sample Purification Beads (Cat No. NEB #E7405) per the manufacturer’s protocols with the total starting RNA concentration of 100ng-1μg. rRNA-depleted RNA libraries were then prepared using NEBNext® Ultra™ II Directional RNA Library Prep (Cat. No. E7765L) with Sample Purification Beads and sequenced on the NovaSeq 6000 platform using an S4 flow cell and a 2x150bp read length kit (Illumina).

#### (ii) Targeted sequencing of SARS-CoV-2

The libraries were prepared using the Illumina COVIDSeq Test kit (Cat No: 20043675, Illumina Inc, USA). Extracted RNA samples were primed by random hexamers for reverse transcription. The complementary DNA (cDNA) products were amplified using ARTIC V3 and ARCTIC V5.3.2 (for December 2023 samples) primer set targeting the entire SARS-CoV-2 genome and human cDNA targets in two different multiplex PCR reactions. The amplified product was later processed for tagmentation, and adapter ligation using IDT for Illumina Nextera DNA Unique Dual Indexes Sets A–D IDT for Illumina-PCR Indexes Sets 1–4 (384 Indexes, Cat no: 20043137, Illumina Inc, USA). Further enrichment and clean-up were performed as per the manufacturer’s instructions.

Pooled libraries were quantified using a qubit 4.0 fluorometer (Invitrogen, USA), and library sizes were analysed using TapeStation 4200 (Agilent, USA). The libraries were normalised to 2 nM and denatured with 0.1 N NaOH.

The denatured libraries were sequenced at appropriate concentrations depending on the sequencing system, which included the HiSeq 2500 system with a Rapid SR flow cell (v2: 1×50 bp), the MiSeq system with a flow cell (2×75 bp), and the NovaSeq 6000 system with an SP flow cell (2×100 bp), as per the manufacturer’s instructions (Illumina Inc.)

### 2.5 Bioinformatics

#### (i) Influenza read detection and subtype assignment

We used two bioinformatic pipelines to detect influenza read detection and subtypes assignments. First pipeline performed reference-based assembly using the RefSeq genomes of influenza viruses (A/B/C). Demultiplexed raw fastq files were processed using Trimmomatic (v.0.39) [16] (LEADING:3 TRAILING:3 SLIDINGWINDOW:4:30 MINLEN:50) to remove Illumina adapters. Potential human reads were removed using Kraken Human database (Kraken v.2.1.3) [17] downloaded using kraken2-build command function. Paired end trimmed reads were re-paired with the repair.sh utility from bbmap (sourceforge.net/projects/bbmap/). Trimmed FASTQs were mapped to all influenza whole genome sequences available in the Influenza Resource Database (IRD) of National Centre for Biotechnology Information (NCBI). Bowtie2 (v.2.2.5) [18] was used to map quality-controlled reads to the influenza database using influenza.fasta file https://ftp.ncbi.nih.gov/genomes/INFLUENZA/influenza.fna (downloaded on 08.04.2023). SAMtools coverage/bedcov [19] was used to calculate the basic stats of the mapped file resulting in read count calculations for each subtype, read length. Bam ‘*validate*’ is used to calculate the mapping rate of each sample. Sample metadata from NCBI was used to annotate likely subtypes and hosts of influenza virus sequences detected in each collecting location, based on the closest match in the NCBI. To avoid false positives resulting from index swapping during capture [20–21], we established a threshold for each virus, consisting of 0.1% of the total reads for that virus in the appropriate pool. This was based on published studies and is consistent with our experiences dealing with dual-indexed sequencing libraries and the HiSeq 2500 platform. Samples were required to have a greater percentage of reads assigned to a particular virus than the percentage of reads assigned to that virus across all batch-specific controls. Read counts were further normalized as reads per million (RPM) total (trimmed) reads. To reduce false-positive virus discovery, an arbitrary acceptance criterion of viruses with a threshold of RPMLJ>LJ1 for a significant metagenomic NGS finding were considered [22].

Second pipeline used the publicly available 981,537 iav_serotype (v0.1.4) software package (https://github.com/mtisza1/influenza_a_serotype). Briefly, paired-end reads were quality filtered using fastp (v0.23.4) with default settings. Reads are aligned to Influenza_A_segment_sequences database v1.1 using minimap23 (v2.28-r1209) and flags “-cx sr --secondary=yes -f 100000” to allow secondary alignments. Average nucleotide identity (ANI) and alignment fraction (AF) were assigned to a particular serotype if the best alignment is exclusive to one serotype and (ANI*AF >= 0.9). Otherwise, reads are assigned as “ambiguous”.

Data processing and visualization was implemented in Rstudio (v.4.2.1) [23], using multiple packages (ggplot2, tidyverse, corrplot, Hmisc).

#### (ii) SARS-CoV-2 variant detection

Wastewater samples consist of a mixture of variants circulating in a population in contrast with clinical samples which might be infected with a single variant. In the first step, the raw reads were aligned with the reference genome of SARS-CoV-2 (MN908947). The processed reads are aligned with the reference genome by BWA Mem [24] and various coverage statistics are taken by SAMtools coverage/bedcov [19]. The alignment was used for a single nucleotide variant (SNV) calling using iVar [25]. The iVar tool was used to trim the primers and generate a table for each sample with mutation frequency data and an adj p-value (Fisher’s test) for altered positions of SARS-CoV-2 from the BAM files. iVar was run with a minimum base quality filter of 20 (default value) using the reference genome of SARS-CoV-2 and the feature file Sars_cov_2.ASM985889v3.101.gff3 from NCBI. For predicting the lineage abundances, a deconvolution matrix was generated using *Freyja* (https://github.com/andersen-lab/Freyja) [26] -a dedicated bioinformatic pipeline for wastewater analysis. From measurements of SNV frequency and sequencing depth at each position in the genome, *Freyja* returns an estimate of the true lineage abundances in the sample. *Frejya* used the UShER tree with WHO designation and *outbreak.info* metadata [27].

## 3. Results

We screened 812 wastewater samples collected from 28 sewershed sites across Bengaluru (Fig. 1) between August 1, 2021, and December 31, 2023. RNA from all target viruses was detected in wastewater. The abundance of SARS-CoV-2 was significantly higher than other target viruses. SARS-CoV-2 was detected in 86% (699/812) of samples. IAV and IBV were detected in 37% (305/812), and 16% (131/812) of the total samples, respectively. However, RSV was detected in 15% (123/812) of samples.

The analysis of influenza virus revealed that IAV always predominated over IBV, reaching a prominent peak in post monsoon season i.e. August to December followed by January and February, while being practically absent the rest of the year. Compared to IBV, RSV showed similar trend to IAV was present in similar quantities (Fig.1A). Furthermore, IAV and RSV positivity and its concentration varied across the sewershed sites (Fig. 2). SARS-CoV-2 was prevalent throughout the study period and there were four main surges were recorded by wastewater genomic data: BA.2.10 surge, BA.2.X surge, XBB surge, and JN.1 surge. However, there was no overlap in SARS-CoV-2 surges and IAV and RSV peaks. In contrast, IAV and RSV exhibited a seasonal pattern whereas SARS-CoV-2 was driven by the emergence of new variants. This was further corroborated by Kendall tau statistics. SARS-CoV-2 RNA was not significantly correlated with any viral target. However, IAV, IBV, and RSV exhibited significant cross-correlation, with tau values ranging from 0.46 (IAV and IBV) to 0.23 (IAV and RSV) and 0.21 (IBV and RSV) (Supplementary Table 2).

**Fig. 2.**
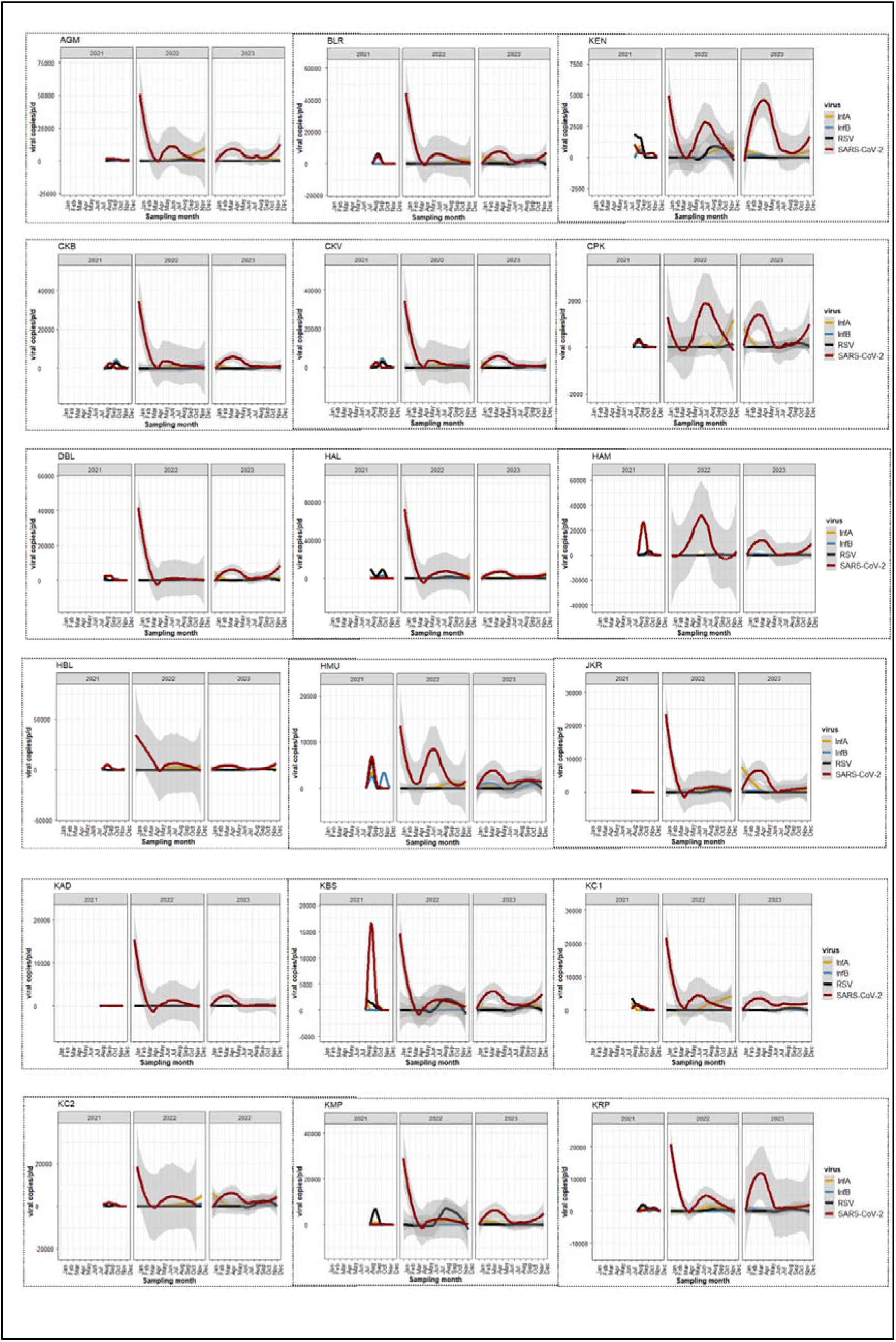
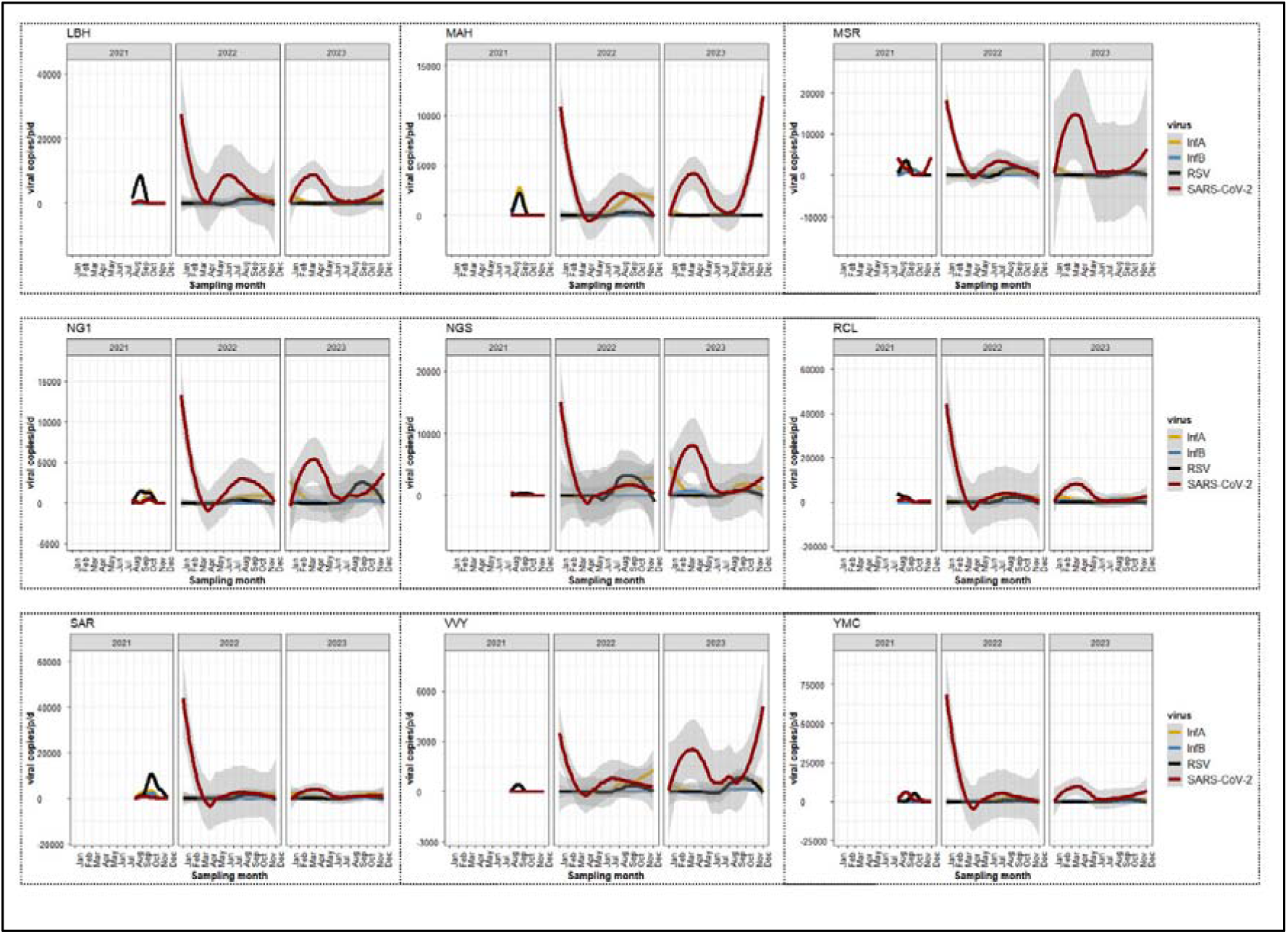
Temporal dynamics of normalized viral load (gene copies/person/day) of SARS-COV-2, IAV, IBV and RSV across sewershed sites.

### 3.1 Influenza virus read abundance and alignment to genome segments

We analysed a subset of 242 wastewater samples from August 2021 to December 2023 and details on total reads, sequencing depth is in Supplementary Table 3 and Supplementary Table 4. Using all influenza virus analysis, the genome coverage ranged between 1.3% to 33%. The IAV reads in wastewater samples ranged from 1 to 86, with 2405 sequences detected and no single sample producing reads for every segment (Supplementary Fig.1). The reads aligned to all eight genome segments and were generally biased towards segment 1 [(PB2=369 (IAV); 6 (IBV); 10 (Inf C)], segment 3 [PA=1702 (IAV); 12 (IBV)], and segment 7 [(M= 137 (IAV); 6 (IBV); 6 (Inf C)] (Fig. 3). The segment 2 [PB1= 19 (IAV); 2 (IBV);16 (Inf C)], segment 4 [HA= 14 (IAV); 11 (IBV); 10 (Inf C)], segment 5 [NP =15 (IAV); 4 (IBC);10 (Inf C)], segment 6 [NA= 2 (IAV); 2 (Inf C)], segment 8 [NS= 4 (IAV); 4 (IBV); 30 (Inf C)] had >20 matches in the database. In addition, influenza C had hemagglutinin-esterase (HE) gene= 4 and P3 gene= 2. The mean read number and coverage were 7⋅6% for the HA segment and 10.9% for the NA segment.

**Fig. 3.**
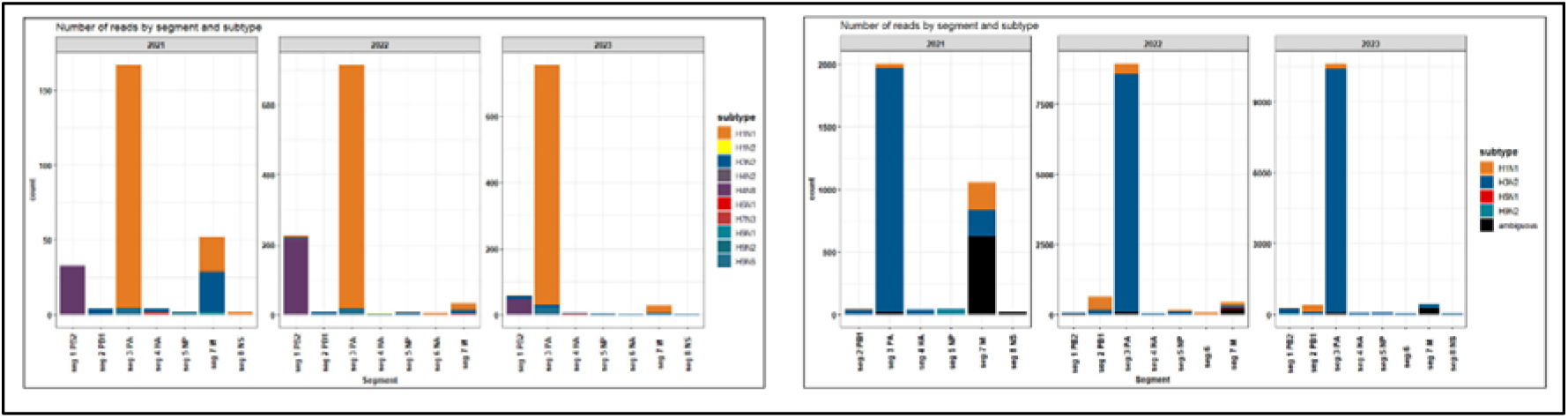
Number of reads amplified and sequenced from wastewater that matched database of influenza virus whole genomes using two bioinformatic pipelines: left: NCBI RefSeq Influenza genomes; right: iav_serotype.

Similarly, iav_serotype pipeline assigned reads to all eight segments of influenza A genome and biased towards segment 3 and segment 7 (Fig. 3). However, iav_serotype software tends to follow a conservative approach that can obscure minority strains, the read alignments were assigned to a particular subtype if the best alignment was exclusive to one subtype and ANI*AF >=0.9. Using iav-serotype, the reads were assigned to seasonal influenza subtypes, H3N2 and H1N1.

### 3.2 Influenza diversity in wastewater

Using RefSeq alignments, wastewater sequences identified ten influenza A subtypes (H1N1, H1N2, H3N2, H4N8, H5N1, H7N3, H9N1, H9N2 and H9N5), one influenza B subtype (Victoria) and influenza C (Fig. 4). H1N1 and H4N8 were the dominant subtypes followed by H3N2.

**Fig. 4.**
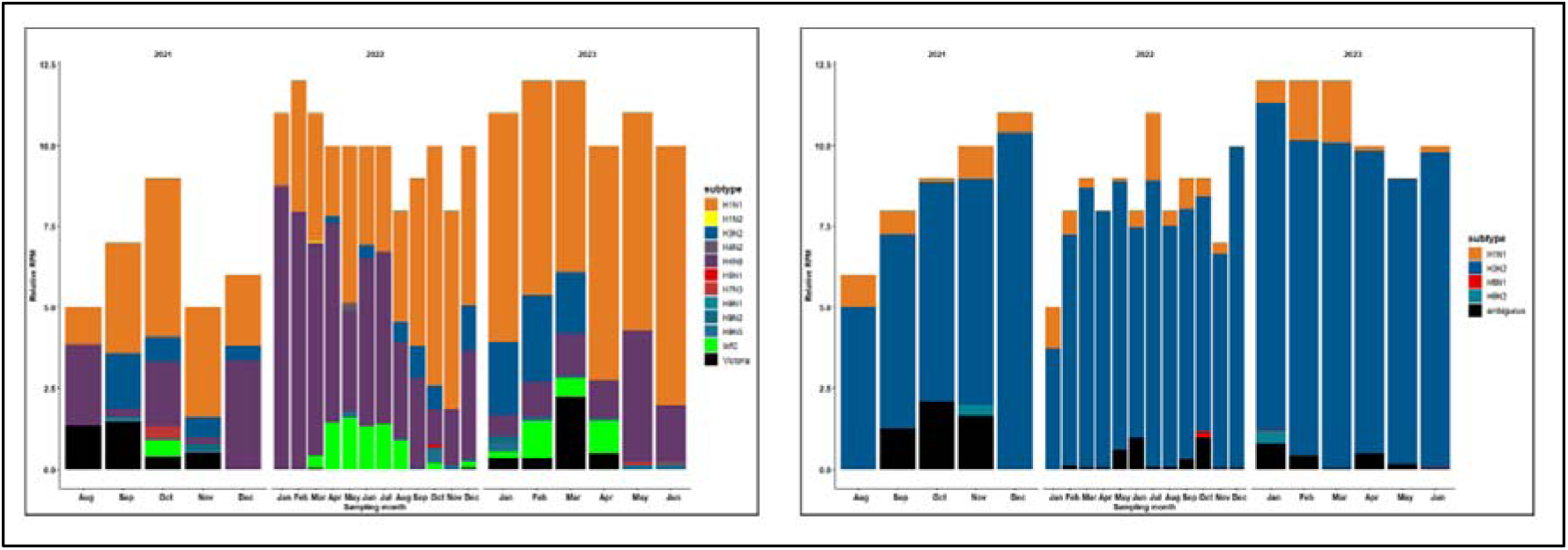
Temporal variation in the relative abundance of influenza subtypes in wastewater in Bengaluru sewage using two bioinformatic pipelines: left: NCBI RefSeq Influenza genomes; right: iav_serotype

Using the iav_serotype pipeline, the reads were aligned to all eight segments representing the metagenomes of influenza subtypes present in a single sample, reflecting the ‘true’ diversity of influenza viruses in wastewater, which otherwise remained undetected using targeted sequencing or RT-PCR approaches. Nonetheless, the patterns in influenza A subtypes H1N1 and H3N2 matched the RT-PCR viral load.

The relative abundance of these subtype showed a variable pattern with space and time (Fig. 1B). Furthermore, the number of reads for human-like influenza subtype (H1N1) was larger than avian-like subtype (H4N8 and H9N5) (Fig. 5).

**Fig. 5.**
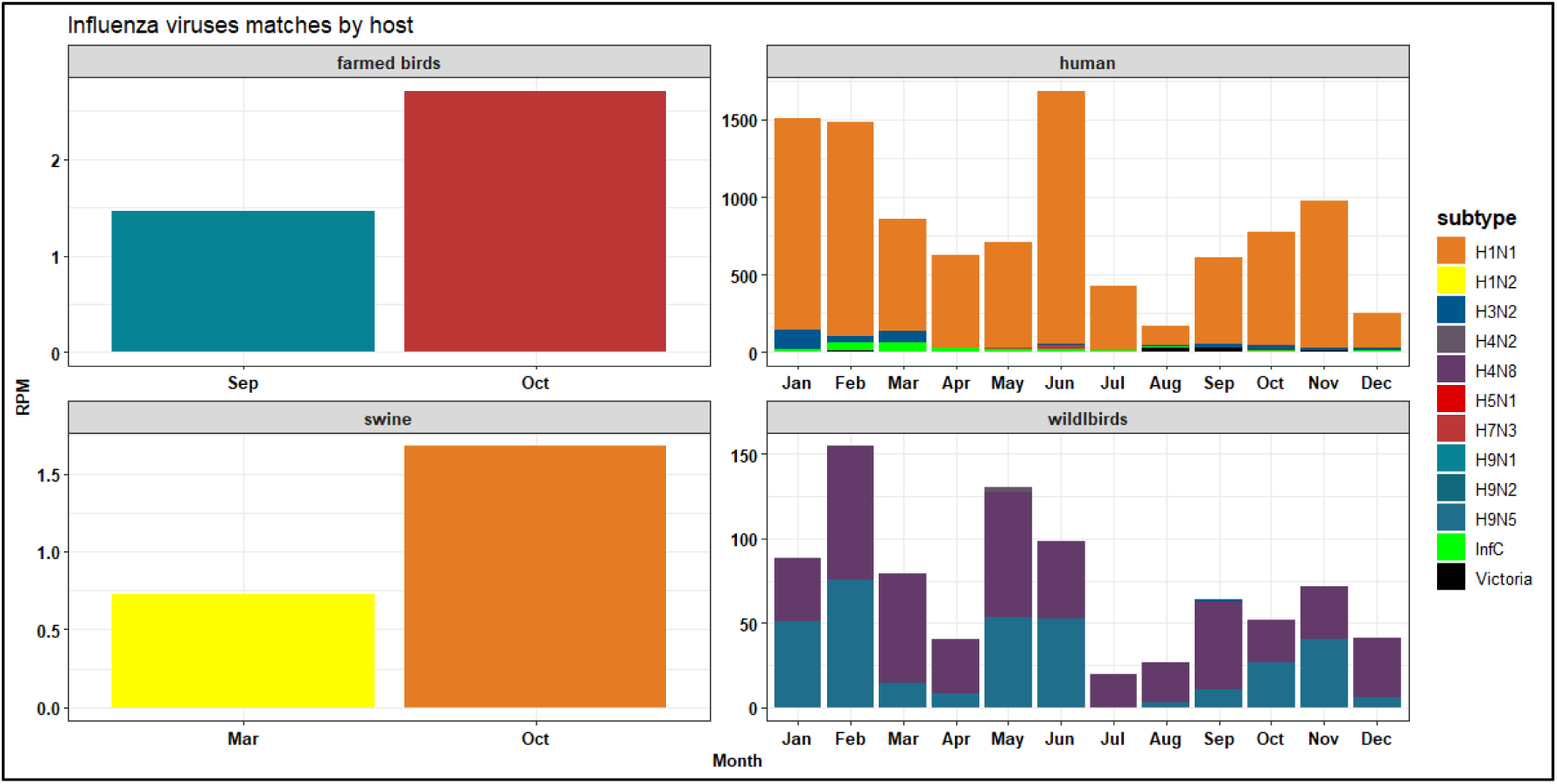
Proportion of unique reads from samples were assigned a similarity to human-like and avian like influenza subtypes based on similarity in NCBI database.

## 4. Discussion

We show that wastewater surveillance is an effective tool for monitoring infectious diseases and understanding the transmission dynamics of respiratory viruses beyond SARS-CoV-2. Wastewater data on the abundance of respiratory viruses in Bengaluru city provided new information which is not available via clinical surveillance and can be used to identify circulating strains associated with poultry and other farmed birds. Using an untargeted metagenomic approach on wastewater samples, we rapidly determined various subtypes of influenza viruses, showing concordance between RT-qPCR and read counts. However, it is important to note that RT-qPCR is optimized to assess infection severity and does not capture signals below a certain threshold, i.e., low viral load. In contrast, Illumina sequencing can achieve accuracies and sensitivities with target hits as low as one read to confirm the presence of a pathogen and has been shown to be highly comparable to commercial viral panels (e.g., Graf *et al*. [28]). The unbiased nature of sequencing allows us to query a theoretically unlimited number of pathogens in parallel, increasing the chance of detecting all pathogens, including human viruses, and resulting in a higher positivity rate. We used sequencing reads from samples with low read counts (samples produced <3 reads per subtype, below our arbitrary significance level of RPM>1). Our results demonstrate that untargeted metagenomics can be used to supplement PCR-based tests and that wastewater metagenomic data can be used to investigate viral etiologies of respiratory infections in low-resource settings in two contexts: (i) to understand what pathogen is driving the seasonal illness patterns at a community level. We revealed a significant diversity IAVs dominated by H1N1, H4N8, and H3N2. Low RSV and IBV viral load resulted in a limited number of reads and the absence of Yamagata/IBV; (ii) to help define testing priorities and vaccination drives. The seasonality of influenza varies within India and often complicates appropriate vaccination recommendations, particularly the timing of vaccination campaigns in tropical regions [29]. We detected a dynamic signal in IAV and IBV in wastewater across sewershed sites. While there is no clinical data from Bengaluru to compare these patterns, wastewater data matches regional trends influenza positivity in Karnataka (Bengaluru Medical College and Research Institute) from October to November 2021 and July to September in 2022 [30]. This data facilitates epidemiologic investigations and studies of vaccine effectiveness. Real-time sequencing of wastewater for influenza will improve surveillance, enable early detection of antiviral drug resistance (reducing indiscriminate antibiotic usage), and inform vaccination strategies.

Changes in human behaviour driven by non-pharmaceutical interventions (e.g., masking, hand washing, reduced movement) during COVID-19 reshaped the evolution and distribution of seasonal influenza viruses [31–32]. Eden *et al*. [33] recorded reduced genetic diversity in RSV during the acute phase of the COVID-19 pandemic and extensive outbreaks in early 2021 upon relaxation of COVID-19 control measures. However, there is limited evidence from India [34] where the decline in influenza incidence is associated with COVID-19-related disruptions, poor health-seeking behavior, and reduced testing due to overburdened health systems, resulting in fewer influenza cases rather than a true reduction in disease transmission [35]. The absence of influenza B/Yamagata further confirms the trend observed elsewhere as a possible extinction during the pandemic. This emphasises the importance of robust and timely surveillance data to inform the selection of vaccine components, and continued surveillance is needed to examine whether these changes will result in sustained seasonal shifts in the post-pandemic years.

Cities offer a heterogenous ever-changing environment, interspersed with pet markets, poultry, and livestock. Often, the blurred rural boundaries and dense populations create potential hotspots for emerging infectious diseases (e.g., severe acute respiratory syndrome (SARS) and avian flu). The presence of avian-like H4N8 and H9N2 highlights the need for a One Health approach to influenza surveillance and the identification of transmission drivers (e.g., increasing human and poultry population numbers, movements, and poor biosafety and biosecurity practices in backyard poultry farms and markets). Tackling this issue warrants multisectoral collaboration between human and veterinary health sectors, with exchange of surveillance data to enable policy.

## 5. Conclusion

We demonstrated that wastewater surveillance can detecting and tracking the SARS-COV-2 infections, variant diversity and shifts in influenza virus transmission dynamics at community level. The viral load captured the COVID-19 infection patterns and distinguish between seasonal influenza dynamics and the SARS-CoV-2 outbreak patterns. Near real-time genomic surveillance is the key to understanding the emerging patterns in viral load and help in defining testing priorities and vaccination drives. This proof-of-concept study can be used in devising guidelines for monitoring human and animal respiratory viruses.

## Data Availability

Data is included in supplementary material. All raw wastewater sequencing data will be available via the NCBI Sequence Read Archive under the BioProject ID PRJNXXX.

## Contributors

FI: conceptualised and designed the study; ND: conducted bioinformatic analysis; SKK: conducted laboratory experiments and sequencing. VS: organised sampling permissions; RM: project management; FI analysed the data and wrote the manuscript. All authors approved the final version of the manuscript.

## Funding

This work has been supported by funding from the Rockefeller Foundation (grant 2021 HTH 018) and the Indian Council of Medical Research grant to (FI) Tata Institute for Genetics and Society and Tata Trusts.

## Data availability

All raw wastewater sequencing data will be available via the NCBI Sequence Read Archive under the BioProject ID PRJNXXX. Consensus sequences from clinical surveillance are all available on GISAID. Freyja is hosted publicly on github (https://github.com/andersen-lab/Freyja) and is available under a BSD-2-Clause License (doi: 10.5281/zenodo.6585067, version 1.3.7). Freyja is accessible as a package via bioconda (https://bioconda.github.io/recipes/freyja/README.html)

## Competing interests

The authors have declared that no competing interests exist.

## Acknowledgments

This work was supported by funding from the Rockefeller Foundation (grant 2021 HTH 018) under the Alliance for Pathogen Surveillance Innovations (APSI)-India, multi-city consortium for wastewater surveillance and the Indian Council of Medical Research grant to (FI) the Tata Institute for Genetics and Society and Tata Trusts. We thank the Bangalore Water Supply and Sewerage Board for providing access to the sewage treatment plants for each sewershed site. We also acknowledge the support of the Director of the National Centre for Biological Sciences (NCBS-TIFR) for facilitating the funding and the NCBS sequencing facility for sequencing the wastewater samples.

## Notes

### Competing Interest Statement

The authors have declared no competing interest.

## References

1. World Health Organization (WHO) Geneva: WHO and UNICEF; 1998. Management of childhood illness in developing countries-rationale for an integrated strategy.

2. Liu, Li et al. National, regional, and state-level all-cause and cause-specific under-5 mortality in India in 2000–15: a systematic analysis with implications for the Sustainable Development Goals. The Lancet Global Health, Volume 7, Issue 6 (2019) e721 − e734.

3. Chotpitayasunondh, T., Fischer, T.K.K., Heraud, J.M.M., et al. Influenza and COVID-19: what does coexistence mean? Influenza Other Respir. Viruses 15 (2021) 407–412.

4. Huang, C., Wang, Y., Li, X., et al. Clinical features of patients infected with 2019 novel coronavirus in Wuhan, China. Lancet 395 (2020) 497.

5. Yang, Jing et al. Co-existence and co-infection of influenza A viruses and coronaviruses: Public health challenges. The Innovation, Volume 3, Issue 5 (2022) 100306.

6. Brammer, L., Budd A., Cox, N., et al. Seasonal and pandemic influenza surveillance considerations for constructing multicomponent systems. Influenza and other Respiratory Viruses 3(2) (2009) 51–58.

7. Krammer F, Smith GJD, Fouchier RAM et al. Influenza. Nat Rev Dis Primers. (2018) 4:3.

8. Bharaj P, Sullender WM, Kabra SK, Mani K, Cherian J, Tyagi V et al. Respiratory viral infections detected by multiplex PCR among pediatric patients with lower respiratory tract infections seen at an urban hospital in Delhi from 2005 to 2007. Virol J (2009) 26(6) :89

9. Yeolekar LR, Damle RG, Kamat AN et al. Respiratory viruses in acute respiratory tract infections in Western India. Indian J Pediatr (2008), 75(4):341–345.

10. Ishtiaq F. Wastewater-based surveillance of vector-borne pathogens. Trends in Parasitology, 40 (2023), 93−95.

11. Lamba S, Sutharsan G et al. SARS-CoV-2 infection dynamics and genomic surveillance to detect variants in wastewater–a longitudinal study in Bengaluru, India. The Lancet Regional Health-Southeast Asia (2023)11.

12. Nataraj A, Mondhe D, Vishwanath S, Ishtiaq F. Metagenomic analysis reveals differential effects of sewage treatment on the microbiome and antibiotic resistome in Bengaluru, India. Water Reuse (2024) 14 (3): 418–433.

13. Isaksoon F, Lundy L, Hedström A, et al. Evaluating the use of alternative normalization approaches on SARS-CoV-2 concentrations in wastewater: Experiences from two catchments in Northern Sweden. Environments. (2022) 9:39.

14. Maurice G. Kendall, “The treatment of ties in ranking problems”, Biometrika Vol. 33, No. 3, pp. 239−251. 1945.

15. Siddle KJ, Eromon P et al. Genomic Analysis of Lassa Virus during an Increase in Cases in Nigeria in 2018 N Engl J Med (2018) 379:1745−1753.

16. Bolger AM, Lohse M, Usadel B. Trimmomatic: a flexible trimmer for Illumina sequence data. Bioinformatics. (2014) 30(15):2114−2120.

17. Lu J, Rincon N, Wood DE, et al. Metagenome analysis using the Kraken software suite. Nat Protoc. (2022)17(12):2815−2839.

18. Langmead B, Trapnell C, Pop M, Salzberg SL. Ultrafast and memory-efficient alignment of short DNA sequences to the human genome. Genome Biol. (2009) 10:1−10.

19. Li H, Handsaker B, Wysoker A, et al. The Sequence alignment/map (SAM) format and SAMtools. Bioinformatics. (2009) 25(16):2078–2079.

20. Kircher M, Heyn P, Kelso J. Addressing challenges in the production and analysis of illumina sequencing data. BMC Genomics (2011)12:382.

21. Kircher M, Sawyer S, Meyer M. Double indexing overcomes inaccuracies in multiplex sequencing on the Illumina platform. Nucleic Acids Res (2012) 40: e3.

22. 22. Van Borm S, Fu Q, Winand R et al. (2020) Evaluation of a commercial exogenous internal process control for diagnostic RNA virus metagenomics from different animal clinical samples. J Virol Methods 283 (2020)113916.

23. RStudio Team. RStudio: Integrated Development for R (Version 4.2.1). Boston, MA: RStudio, PBC; 2023. Available at: http://www.rstudio.com/

24. Li H. Aligning sequence reads, clone sequences and assembly con*gs with BWA-MEM. figshare. Poster 10.6084/m9.figshare.963153.v1; 2014.

25. Grubaugh ND, Gangavarapu K, Quick J, et al. An amplicon-based sequencing framework for accurately measuring intrahost virus diversity using PrimalSeq and iVar. Genome Biol. (2019) 20:8.

26. Karthikeyan S, Levy JI, De Hoff P, et al. Wastewater sequencing reveals early cryptic SARS-CoV-2 variant transmission. Nature (2022) 609:101–108.

27. Tsueng G, Mullen JL, Alkuzweny M, et al. Outbreak Info Research Library: A Standardized, Searchable Platform To Discover and Explore COVID-19 Resources. 2021.

28. Graf EH, Simmon KE, Tardif KD et al. Unbiased Detection of Respiratory Viruses by Use of RNA Sequencing-Based Metagenomics: a Systematic Comparison to a Commercial PCR Panel. J Clin Microbiol (2016) 54.

29. Koul PA, Koul HP. Redefining the influenza equator. The Lancet Global Health (2022) Volume 10, Issue 10, e1388.

30. Potdar V, Vijay N, Mukhopadhyay L et al. Pan-India influenza-like illness (ILI) and Severe acute respiratory infection (SARI) surveillance: epidemiological, clinical and genomic analysis. Front. Public Health (2023) 11:1218292.

31. Chow, E.J., Uyeki, T.M. & Chu, H.Y. The effects of the COVID-19 pandemic on community respiratory virus activity. Nat Rev Microbiol 21 (2023) 195–210.

32. Chen et al. COVID-19 pandemic interventions reshaped the global dispersal of seasonal influenza viruses Science 386, 639 (2024).

33. Eden, JS., Sikazwe, C., Xie, R. et al. Off-season RSV epidemics in Australia after easing of COVID-19 restrictions. Nat Commun 13, 2884 (2022).

34. Haider, M.; Parvaiz, A.K. Negligible circulation of influenza in COVID times in Northern India. Lung India (2021) 38:401–402.

35. Jayaram A., Jagadesh A, Kumar AMV et al. Trends in influenza infections in three states of India from 2015–2021: Has there been a change during COVID-19 pandemic? Trop. Med. Infect. Dis. (2022) 7, 110.

